# Islamic Perspective on Nursing and the Philosophy of Science

**DOI:** 10.1101/2022.04.28.22274408

**Authors:** Atyanti Isworo

## Abstract

Although nursing theory developed in the modern Western scientific tradition has universal aspects, it is important to consider the different philosophies, beliefs and cultures of each different society to enrich the perspective and practice of nursing that is more sensitive to the patient’s background. As supported by the Transcultural Nursing Theory, in treating patients a nurse needs to pay attention to and respect the cultural preferences and beliefs of the patient. In this context, this article wants to contribute by describing the Islamic perspective on nursing in the context of nursing philosophy. Methods: This literature review uses articles published from 2018-2022 with source from databased ScienceDirect, Scopus, Sage Journals, EBSCO-Host, Springer Link, and ProQuest. The keywords used are Islamic, perspective, nursing, caring. Article selection follows “PRISMA” flow. Results: This systematic review have a total of 8 articles meet the criteria for analysis. This article has relevance both in Muslim-majority countries such as Indonesia, Malaysia and Middle Eastern countries, as well as countries where Muslims are a minority. We found 8 articles presents a number of opinions or research results that examine nursing issues and those related to it from an Islamic perspective. From a number of opinions and research findings studied, it can be concluded that although it can be said to offer universal core values, the Islamic perspective has fundamental differences with Western nursing philosophy or theory, especially in terms of the source of knowledge, the basic view of humans, and the importance of spirituality in nursing.

## Introduction

Although nursing theory developed in the modern Western scientific tradition has universal aspects, it is important to consider the different philosophies, beliefs and cultures of different society to enrich the perspective and practice of nursing that is more sensitive to the patient’s background. As supported by the Transcultural Nursing Theory, in treating patients a nurse needs to pay attention to and respect the cultural preferences and beliefs of the patient. For this reason, nurses must have cultural competence in order to be able to provide optimal care according to the needs and culture of the patient (Maier-Lorentz 2008: 37). Nursing that pays more attention to cultural diversity means bringing together an emic perspective (local knowledge and way of life) with an etic perspective (knowledge from a professional point of view) (Leininger 1999). In this context, this article wants to contribute by describing the Islamic perspective on nursing in the context of nursing philosophy.

The urgency of discussing the Islamic perspective on nursing includes both theoretical and practical interests. Practically, developing an Islamic perspective on nursing can help nurses to provide care that is more in line with the religious beliefs and practices of a Muslim patient. By developing sensitivity to the patient’s religious and cultural diversity, it is hoped that a nurse can develop their nursing knowledge and skills (Ott, Al-Khadhuri & Al-Junaibi 2003). By paying attention to the needs of patients based on their cultural and religious backgrounds, a nurse will not a priori assume that all patients have needs or believe in the same prohibitions (Giger 2017). This practical urgency is especially relevant to nursing practice in Muslim-majority countries, such as Indonesia or Malaysia, as well as in countries where Muslims are a minority (Blankinship 2018).

To get a description of the Islamic perspective on nursing, this article will not dig independently into authoritative sources in Islam, the Koran and the traditions of the Prophet Muhammad, nor will it formulate based on the opinions of Muslim philosophers or religious scholars. Because the purpose of this article has basically been done by experts either directly or indirectly, this article will describe and discuss their opinions or research results. The discussion on the views of the experts was carried out in the context of three important aspects in the philosophy of science, i.e. epistemology, ontology and axiology. In the ontology aspect, the focus of the discussion is on the concept of humans. As mentioned by Benner and Wrubel (1989), in Steven Edwards (2001), assumptions about people are essential in nursing theory. While the axiological aspect means the practical consequences of the Islamic perspective in the epistemological and ontological aspects.

## Method

### METHOD

#### Eligibility Criteria

Determination of the article criteria used the PICOS framework search strategy (Population, Intervention, Comparison, Outcome, Study type) according to the inclusion criteria and exclusion criteria that have been determined as in the table below.

| Criteria | Inclusion | Exclusion |
| --- | --- | --- |
| *Population* | Human | No exclusion criteria |
| *Intervention* | Caring | No exclusion criteria |
| *Comparison* | No comparison | No exclusion criteria |
| *Outcome* | Islamic and Western<br>Perspective | Does not discuss the caring<br>from Islamic or western<br>perspective |

The search literature by specific keywords. Keywords used (“islamic” OR “moslem”) AND (“caring”) AND (“perspective” OR “philosophy”) AND (“nursing”). Initial search resulted started by searching the ScienceDirect database obataining 239 articles, Scopus obtaining 11 articles, Sage Journals obtaining 0 article, EBSCOhost resulting 128 articles, ProQuest resulting 0 article, Springer Link obtaining 0 article. The total titles obtained from this search amounted to 378 articles, which were then entered into the Web Importer (Mendeley) to eliminate duplicate articles. After that, the screening stage was carried out on the titles and abstracts of articles that did not match the research topic. After going through the screening stage, then an assessment of the feasibility of the article is carried out based on the completeness of the complete downloadable article. After that is the screening stage based on the inclusion and exclusion criteria that have been set in the study so that 8 articles are found that are deemed to meet the criterissa for further analysis according to the established theme. A summary of the articles that have been analyzed is listed in the PRISMA (Preferred Reporting Item for Systematic reviews and Meta-Analyses) diagram below:

**Figure 1.**
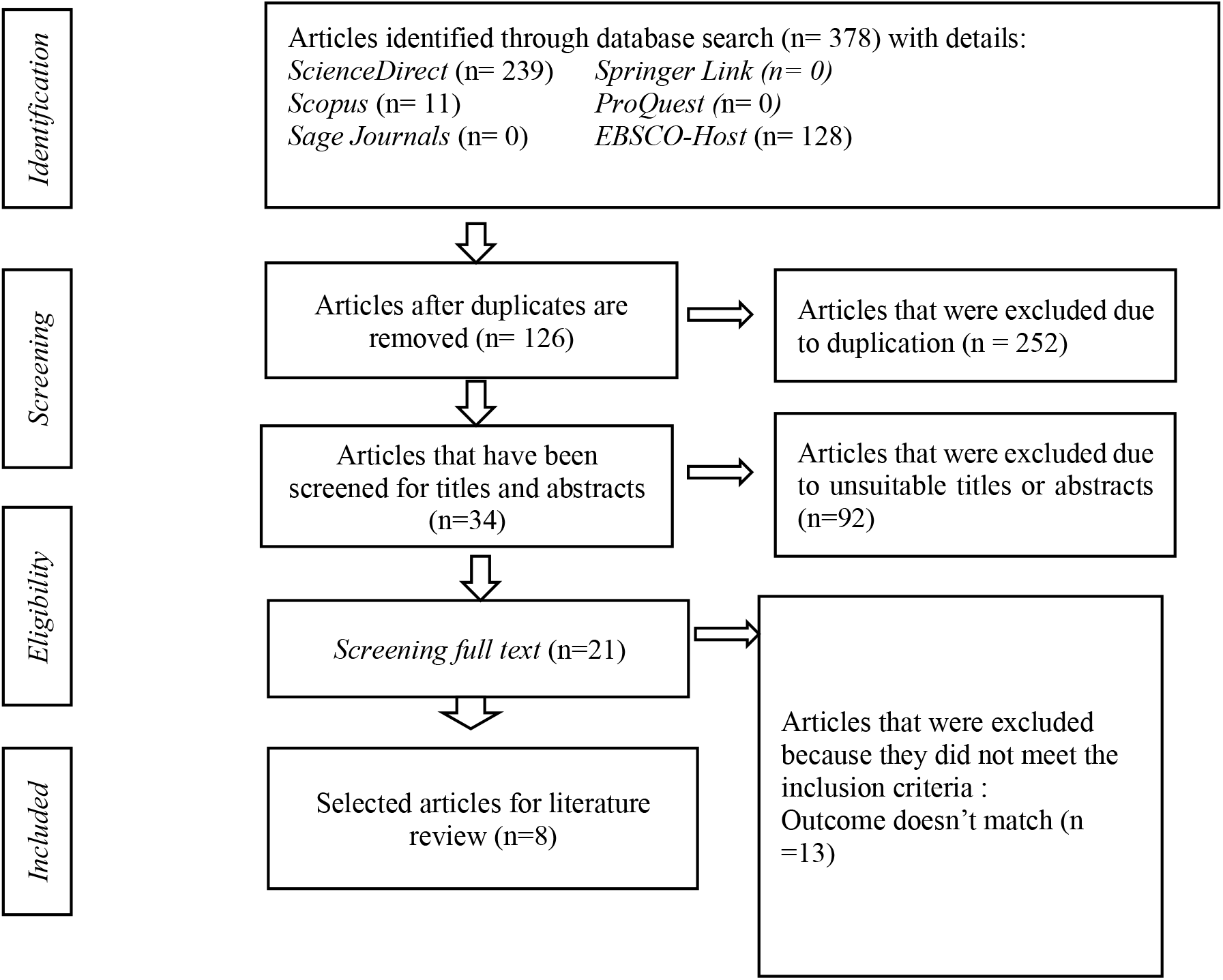
Flowchart PRISMA.

## Result

### Rasool: Caring in Islamic Perspective

According to Rasool (2000) the principles and practices of caring according to Islam are based on God’s revelation. In Islam, caring is an expression of human obedience to God. God expects human beings to love the weak and those who suffers. By doing that, then he is considered to be doing actions that include promoting virtue and preventing vice. Thus, the ethics of caring is rooted in Islamic ethical principles that aim to preserve life, reduce suffering, enjoin good and prevent vice.

Rasool also emphasizes the spiritual aspect by stating that spiritual care occupies an important position in Islam. In fact, according to him, it is very strange if Muslims get treatment by ignoring spiritual care. Although this spiritual aspect is important, the concept of caring in Islam includes compassion for anyone, regardless of their religious background, ethnicity, social status or wealth. Even if Islam opposes alcoholism or homosexual behavior, a Muslim nurse must still provide care for them.

### Barolia & Karmaliani: Theory of caring

Rubina Barolia and Rozina Karmaliani (2008) propose a theory of caring in nursing according to an Islamic perspective which they call an interactive model. They emphasize that the concept of caring is an integral part of Islamic theology. According to the interactive model, balancing the five dimensions of the human personality is important in providing nursing care. The five dimensions are physical, ethical, ideological, spiritual, and intellectual. If a nurse is able to balance the five dimensions he/she will produce caring behavior and actions.

The physical dimension has three important themes, i.e. pain relief, piety (cleanliness) and prevention. The pain in question is not only physical, but also psychological. In Islamic nursing theory, both forms of illness need equal attention. Cleanliness also includes two aspects, i.e. cleanliness/physical piety and mind/heart. The ethical dimension refers to a decision-making process based on principles such as honesty, fairness, and equality to do good for humanity. The ethical dimension also regulates a person’s rights which are regulated in Islam, especially the right to get treatment if sick.

The ideological dimension has three themes that shape the perception of the importance of balance in Islam. The three are obligations to God, obligations to mankind and obligations to oneself. Nursing in Islam requires people to maintain a balance between the practices of the three. The spiritual dimension relates to self-satisfaction obtained by growing affection, building relationships with others, and empathizing with others. The intellectual dimension emphasizes the importance of seeking knowledge in the nursing process. Carrying out research and development of knowledge relevant to the nursing process is basically a command given by God in the Quran.

Barolia and Karmaliani argue that caring in an Islamic perspective basically emphasizes universal nursing moral values which are also shared by other religions and non-religious perspectives. In other words, although there are few concepts related to spirituality in the Islamic perspective that differ philosophically from some experts who argue in the Western tradition, the dimensions of caring in the Islamic perspective have similarities with what was conveyed by some Western nursing experts.

### Lovering: The Crescent of Care Model

The Crescent Care of Model (COCM) was proposed by Sandra Lovering (2012) as a guidance the care of Arab Muslim patients. According to this model, the aim of care is to restore health condition of the patient, yet the focus of care is the patient and family due to family occupies significant position in Arabic society. However, even though the model was developed based on Arabic culture, it is also relevant for other Muslim societies such as those living in Asia that consider the significant value of family. Professional nursing care of this model consists of five components, namely spiritual care, cultural care, psychosocial care, interpersonal care, and clinical care.

Spiritual care which is an action to meet the spiritual needs of patients and their families is a key to nursing actions. Actions included in spiritual care are starting from facilitating patients to worship, ensuring the availability of the Quran to be read to saying ‘*bismillah*’ before starting any procedures. Cultural care is respect for the values, beliefs and traditions of patients and families. For example, the use of traditional medicines or religious healers should also be accommodated in patient care. Psychosocial care is an attempt to determine the family structure so that it can be identified who makes decisions in the family and the position of the patient in the family. Nurses must work closely with the patient’s family because family support can help patients reduce stress and anxiety. Interpersonal care is concerned with the patient-nurse relationship, including patterns of verbal and non-verbal communication. Respect for the values of modesty, especially relations between different genders, such as eye contact and physical touch, is important. Clinical care not only includes the knowledge and skills needed to carry out nursing practice, but also involves the family in patient care and accommodation for the needs of patients being treated during the month of *Ramadan*.

### Sadat Hoseini et al.: Nursing according to the scripture

Sadat Hoseini et al (2013) formulate the concept of nursing in an Islamic perspective by referring to the relevant verses of the Quran. Based on the interpretation of the relevant Quranic verses, they argue that according to Islam nursing is an act of guiding someone to find a solution (remedy) with feminine criteria. This is related to the female character (mother) who always tries to find solutions and foster healing. In implementing the healing methods he has learned based on love, compassion and kindness. Although nursing is considered to be closely related to female character, this does not mean that nursing as a profession is only exclusive to women.

In Islam, the pivot of nursing is the patient, not the nurse. Health is seen as an individual’s internal affair. Nurses will only be involved when a person is unable to care for himself or herself or is dealing with her own problems. The nurse is only the party who is looking for a solution or remedy for the problem. Some of the characteristics of nursing according to the Quran and its interpretation: 1. Nursing is a caregiver who seeks medicine for all aspects of other people, therefore they’re higher than caring. They guide others in need and the unknowing. 2. Nursing is patient-centred and is carried out at the open or closed request of the patient. 3. Nursing has a character like motherhood, but is not a special profession for women. 4. The nursing process is the development of nursing values and capabilities that contribute to self-growth. If someone needs the help of a nurse to get healing then the role of a nurse is like the role of a mother who educates with kindness and enthusiasm (Sadat Hoseini et al 2013).

#### Alimohammadi et al.: Human concept and nursing

Humans occupy an important position in nursing. Therefore Alimohammadi et al. (2014) attempted to reconstruct the concept of human in Islamic thought that might be useful in treating Muslim patients. They argue that humans in the Islamic perspective are a unified reality and a created unitary being has two aspects: material (body or person) and immaterial (soul or spirit). The soul or spirit is joined to the body because of that they appear as a single entity that forms a system of existence.

In their view, human character is manifested in four characteristics/attributes: 1. The cognitive dimension that refers to the ability to collect, process, remember, exchange information, and make rational decisions; 2. The dimensions of emotion that are typical of humans consist of a number of characters: affection and sympathy, mutual understanding and interaction, helping the weak, self-sacrifice. 3. The social dimension that refers to self-actualization that cannot be done alone since humans need cooperation, a social environment, and a civilized place to live. 4. The most important dimension of human attributes is the spiritual dimension. In Islam, spirituality cannot be separated from religiosity because spirituality becomes unimportant if it is separated from religious thoughts and actions. Therefore, spirituality is not only a matter of a tendency to perfection, a search for truth, a tendency to ethics and aesthetics, but also a natural innate desire to worship God.

In addition to understanding the attributes possessed by humans, Islam also introduces boundaries that distinguish humans from other creations, namely humans as representatives of God on earth; human as a noble creature; inclination to perfection; and have the ability to choose. With its distinctive attributes and values that distinguish it from other creations, in Islamic thought, humans are seen as holistic beings. According to Alimohammadi et al. the concept of human in Islam can be compared with other concepts in nursing studies, but at the same time it is different because of its uniqueness.

According to Alimohammadi et al. Islam rejects the dualism perspective in seeing humans as understood by some experts. Meanwhile, the Islamic perspective on humans is also different from the reciprocal interaction worldview; this perspective is considered by them to ignore the spiritual aspect and do not see humans as a single entity that has material and immaterial aspects that interact as a unified whole. Meanwhile, the Islamic perspective is also different from the simoultaneous action worldview; this perspective is considered to have a hidden dualism view, especially in seeing the relationship between humans and the environment.

#### Alharbi & Al Hadid: Compassion in Islamic perspetive

In their article, Alharbi & Al Hadid (2018) propose the concept of compassion from an Islamic perspective. According to them, compassion is an important concept in nursing because it is considered to increase patient satisfaction with the health services provided. Meanwhile, discussions about the concept of compassion according to Islam are still rare compared to similar discussions in Western and Buddhist perspectives. According to them, compassion represents the main spirit of Islamic teachings because it is reflected in a number of words that are often mentioned after the main concepts in Islam, namely *tawhid* (unity of God) and *risalah* (messengership of the prophet); they are mercy (*rahma*), benevolence (*ihsan*) and *adl* (fair and justice). God commands humans through the example given by the Prophet Muhammad to give sympathy and care to all creatures (living and non-living), especially the weak, sick, poor, old people, orphans and the oppressed.

The consequence of the concept of compassion mentioned above to the world of health, especially nursing, is that a nurse must show compassion to those they care for regardless of their religious background or ethnicity. Caring for patients with compassion also means carrying out professional and competent care so that they receive quality, fair and equal treatment. Being a compassionate nurse also means making patients in need of care the focus of attention and, therefore, not intentionally harming them or delaying care.

#### Sadat Hoseini: Islamic nursing conceptual structure

Sadat Hoseini (2019) offers a conceptual framework of nursing in an Islamic perspective which consists of four multidimensional concepts that are related to one another: human, health, environment, and nursing. Humans are understood as creatures that have five dimensions, body, soul, human nature, instincts and Fetrate. Fetrate is the human tendency to acknowledge the existence of God who created the universe. The four dimensions are interconnected with each other. Body and soul are interconnected. The body is related to instinct and human nature, while the soul is related to instinct and Fetrate.

Health consists of five dimensions, health, disease, intellectual health, transcendence, and ultimate wellness (*qalbe selime*). The life process will be manifested in the dichotomy of wellness and disease that will lead to intellectual health. Intellectual health which is understood as the ability to make choices and design the process of life based on wisdom and intellect will then lead to transcendence. Transcendence which is formed from three elements, namely knowledge and understanding of God and life after death, attitudes about God, and behavior that is always based on God’s word and spreads goodness will bring a person to ultimate wellness. Meanwhile, the environment consists of two kinds, namely the natural and the social environment which are interconnected.

Nursing aims to find solutions/medicines for problems both body and soul faced by people in need in order to achieve qalbe salim. An understanding of the concept of human and health affects the nursing process. MFor example, in the assessment stage, a nurse must conduct a holistic assessment that includes the five human dimensions. Likewise in the solution stage which includes basic care, educational remedies, behavioral remedies, ethical remedies, spiritual remedies, and self-management. In conclusion, humans, health, environment and nursing are connected to each other where nurses help humans utilize the natural and social environment to achieve qalbe salim.

#### Almukhaini, Goldberg & Watson: Islamic philosophy and nursing theory

In their article, Almukhaini, Goldberg and Watson (2020) compare between Islamic perspectives, especially those developed by Barolia and Karmaliani above, and Jean Watson on caring. They concluded that there were a number of similarities between the two. KBoth of them emphasize that care must start from oneself and then extend to other people, the community and the universe in general, based on a holistically integrated approach covering all dimensions of human beings. In other words, both see humans holistically which includes both material and immaterial aspects of human beings. In addition, both also emphasize the importance of ethical and moral concepts such as hope, love and compassion as well as mutually beneficial relationships in nursing practice so that the goals of care, i.e. self-transcendence, achievement of self-actualization and recognition of human potential, can be achieved.

The basic difference between the two lies in the epistemological aspect; Watson builds her theory based on existentialismt and phenomenology, while what Barolia and Karmaliani propose is based on Islamic theology. In the first, human excellence and potential can be achieved by making use of one’s own experiences and interactions with others. While in the second, the role of religious beliefs and practices also occupies an important position. Carrying out obligations to God and following the model exemplified by the Prophet Muhammad is essential to realize human potential.

## Discussion

Epistemologically, it appears that the Islamic perspective is unique compared to the modern Western approach to nursing. As a perspective that is built on religious teachings, the Islamic view of nursing is derived from one of the revelations of God in the Quran and the traditions brought by the Prophet Muhammad. Sadat Hoseini et al. shows that there are verses in the Quran that use words which in modern terminology are close to the term nursing. With an understanding of nursing that comes from God’s revelation, Sadat Hoseini et al. shows that the concept of nursing in Islam has similarities and differences with what has been developed in the modern Western nursing tradition. Likewise with Almukhaini, Goldberg and Watson who show the basic differences between Islamic perspectives Western (Watson) perspectives on caring, one of which lies in the source of inspiration. The existence of a wedge with the concept developed by experts in the Western world shows that there is an aspect of universality in the Islamic perspective on nursing.

The epistemological differences between the Islamic perspective and modern nursing lead to important ontological differences. In the study of nursing philosophy, one of the important issues in ontology is how humans are understood.In explaining the Islamic perspective on humans which has implications for nursing theory, experts agree that humans have a number of dimensions. The difference between them is in the details of the human dimension. What they have in common is that they argue that human beings have physical/body and soul/spiritual dimensions. Humans are both material and immaterial beings. This view is not typical to Islam. Other religions or some modern Western nursing experts also have a similar view. However, for experts like Alimohammadi et al. the relationship between the material and immaterial dimensions is not seen in the lens of dualism. Both are seen as two dimensions of a single entity that interacts with each other continuously. The Islamic perspective that sees humans holistically can be said to be unique to Islam but is also relevant to the core values of universal humanity.

In his explanation of the interactive model of caring theory, Barolia & Karmaliani argue that caring in an Islamic perspective basically emphasizes universal human values that are also shared by other religions or non-religious perspectives. That is, the five dimensions of the human personality which are the basis for nursing care according to the interactive model, i.e. physical, ethical, ideological, spiritual, and intellectual, are basically recognized by other religions. The offer of caring theory in an interactive model that emphasizes the balance of the five dimensions of the human personality is also considered relevant to be applied to non-Muslim patients. By recognizing the importance of the spiritual aspect in the nursing process, the patient’s religion or beliefs that are not Muslim will still be respected. Rasool and Alharbi & Al Hadid also emphasized that the axiological consequence of the importance of spiritualism in nursing practice from an Islamic perspective also means that compassion should be given to everyone. Even if a patient practices a lifestyle that is contrary to Islam, a Muslim nurse is still obligated to treat him with compassion.

Although there is an acknowledgment of the universal values of Islam in the matter of nursing, some other experts underline the importance of the spiritual aspect in Islamic nursing practice. Experts such as Barolia & Karmaliani do state that caring theory in Islam emphasizes the balance of the five dimensions of the human personality. While Sadat Hoseini et al. assume that mental care (spiritual) is more important than physical care. If we take a position as supported by Hoseini et al. then the Islamic perspective in this case is different from Western nursing experts who prioritize physical or at least see the position of the soul (spiritual) and physical aspects equally. Apart from these differences, the importance of the spiritual aspect in Islamic nursing also has other axiological consequences. Hoseini argues that the spiritual aspect is one of the things that is considered in the holistic treatment process, starting from the assessment stage which covers all human dimensions to the solution stage which includes spiritual treatment. The spiritual aspect is considered starting from assessment to treatment because the concept of health in Islam is not only physical health but also health that has been transcended (ultimate wellness), which is awareness of God which is manifested in attitudes and behavior that always refers to divine inspiration. Lovering also contends that spiritual care is essential in her the Crescent of Care model

## Conclusion

From a number of opinions and research findings studied, it can be concluded that although it can be said to offer universal human values, the Islamic perspective on nursing has fundamental differences with Western nursing philosophy or theory, especially in terms of the basic view of humans and the importance of spirituality in nursing. Epistemologically, the basic difference between the two is rooted in recognized sources of knowledge. The experts above assert that nursing in Islam is built on Islamic theology which is based on the Quran and the example of the Prophet Muhammad. Ontologically, the concept of human, and its relation to health, the environment and nursing, according to Islam has its own uniqueness, although there are intersections with a number of concepts built in the tradition of modern Western knowledge.

The challenge for developing an Islamic perspective on nursing is to examine the concepts it constructs in nursing research and practices. Moreover, the above experts said that nursing in Islam is not meant to be exclusive to patients who are Muslim. Ontological human understanding must have axiological consequences that can be studied; understanding of humans as a single entity that has material and immaterial aspects must be further derived both in research and nursing practices. Axiologically, it must be shown that the spiritual care intended in Islamic nursing contributes to better healing for the patient than if it was completely ignored.

## Data Availability

All data produced in the present work are contained in the manuscript

